# Critical success factors for high routine immunization performance: A case study of Nepal

**DOI:** 10.1101/2022.01.28.22270023

**Authors:** Kyra A Hester, Zoe Sakas, Anna S. Ellis, Anindya Sekhar Bose, Roopa Darwar, Jhalak Gautam, Chandni Jaishwal, Hanleigh James, Pinar Keskinocak, Dima Nazzal, Emily Awino Ogutu, Katie Rodriguez, Francisco Castillo Zunino, Sameer Dixit, Robert A. Bednarczyk, Matthew C. Freeman, the Vaccine Exemplars Research Consortium

**Author notes:** Correspondence to; 1518 Clifton Road NE, Atlanta, GA, 30322. **Contributions:** KAH, AE, SD, RAB, MCF: project conceptualization and methodology; KAH, ZS, ASE, KR, ASB, SD, RAB, MCF: investigation and data curation; KAH, ZS, ASE, RD, CJ, HJ, PK, KR, DN, EAO, FCZ, RAB, ASB, SD, MCF: formal analysis; KAH, ZS, RD, ASE, KR, CJ, HL, EAO, FCZ: writing - original; KAH, ASE, ZS, FCZ, EAO, RAB, MCF: writing – review and editing; all authors provided approval of the submitted version. **Consortium Contributions:** NB, MRD, CE, KI, WO, MR: project conceptualization; RMR, DS, BPS, NS: investigation and data curation; SB, SD, MRD, CE, KRI, RMR, BPM, WO, NB, BPM, NRR, SR, SD, SPS, NS: formal analysis; NB, MRD, CE, KRI, BPM, MRR, WO, SR: writing – review and editing; all authors provided approval of the submitted version.

## Abstract

**Introduction:** The essential components of a vaccine delivery system are well-documented, but robust evidence on how and why the related processes and implementation strategies drive catalytic improvements in vaccination coverage are not well established. To address this gap, we identified critical success factors that may have led to substantial improvements in routine childhood immunization coverage in Nepal from 2000 through 2019.

**Methods:** We identified Nepal as an exemplar in the delivery of early childhood vaccines through analysis of DTP1 and DTP3 coverage data. Through interviews and focus group discussions at the national, regional, district, health post, and community level, we investigated factors that contributed to high and sustained vaccine coverage. We conducted a thematic analysis through application of implementation science frameworks to determine critical success factors. We triangulated these findings with quantitative analyses using publicly available data.

**Results:** The following success factors emerged: 1) Codification of health as a human right, along with other vaccine-specific legislation, ensured the stability of vaccination programming; 2) National and multi-national partnerships supported information sharing, division of labor, and mutual capacity building; 3) Pro-vaccine messaging through various mediums, which was tailored to local needs, generated public awareness; 4) Female Community Health Volunteers educated community members as trusted and compassionate neighbors; and 5) Cultural values fostered collective responsibility and community ownership of vaccine coverage.

**Conclusion:** This case study of Nepal suggests that the success of its national immunization program relied on the engagement and understanding of the beneficiaries. The immunization program was supported by consistent and reliable commitment, collaboration, awareness, and collective responsibility between the government, community, and partners. These networks are strengthened through a collective dedication to vaccination programming and a universal belief in health as a human right.

## 1. Introduction

Vaccination is one of the most influential public health interventions of the last century, preventing an estimated 2-3 million deaths annually [1-3]. The Global Vaccine Action Plan (GVAP) targeted at least 90% national coverage of the third dose of diphtheria, tetanus, pertussis vaccine (DTP3) among 1-year-old children, a globally recognized proxy for vaccination system performance, and at least 80% DTP3 coverage for subnational levels [4]. By 2018, only 95 of the 193 World Health Organization (WHO) Member States achieved the GVAP targets [5]. From 2000 to 2016, the South-East Asia Region (SEAR) consistently reported one of the lowest DTP3 coverage rates with an average rate of 78%; however, DTP3 coverage in the SEAR has significantly increased and averaged 91% as of 2019 [6]. Moreover, Nepal has outperformed its peers by increasing its DTP3 coverage from 74% and 93% between 2000 and 2019, as illustrated in Figure 3. Examining Nepal’s success provides an opportunity to identify and describe critical factors for effective vaccine systems.

The essential components of an effective vaccine delivery system are well established and include strong leadership and governance, healthcare financing, human resources, supply chain, and information systems [7]. Determinants of vaccine coverage include intent to vaccinate, community access, and health facility capacity [8]. However, evidence is lacking on how policies and implementation strategies are operationalized - through strong governance and financing mechanisms - to drive and sustain catalytic changes in coverage. The impact of contextual factors on service delivery has seldom been reported [7].

The purpose of this study was to identify critical success factors that contributed to exemplary growth in childhood routine immunization coverage in Nepal. Findings from this research may identify transferable lessons and support actionable recommendations to improve national immunization coverage in other settings [9]. We applied a positive deviance approach through investigating vaccine delivery systems in exemplary countries [10]. Here we describe the design, adaptation, and implementation of successful policies and programs in Nepal.

## 2. Materials and methods

We applied a mixed-methods CASE study design to explore critical success factors of the Nepali vaccine delivery system. This study was nested within the Exemplars in Vaccine Delivery project to identify components that supported immunization coverage improvements across three exemplary countries – Zambia, Nepal, and Senegal [10]. Based on available data and logistical considerations, the analytic periods varied: Data from 2000-2016 was used for site selection; data from 2000-2019 for quantitative analysis; and qualitative data collection and analysis occurred between 2000-2019. Topic guides utilized coverage graphs from 2000-2018, but key informants also spoke to recent programming. Therefore, the inclusive study period is 2000-2019.

### 2.1. Conceptual Model

Prior to data collection, we developed a conceptual model (Figure 1) to organize factors impacting childhood vaccine coverage globally. This model was based on the work of Phillips et al. and LaFond et al. alongside a broader review of the vaccine confidence and coverage literature [8, 11].

**Figure 1.**
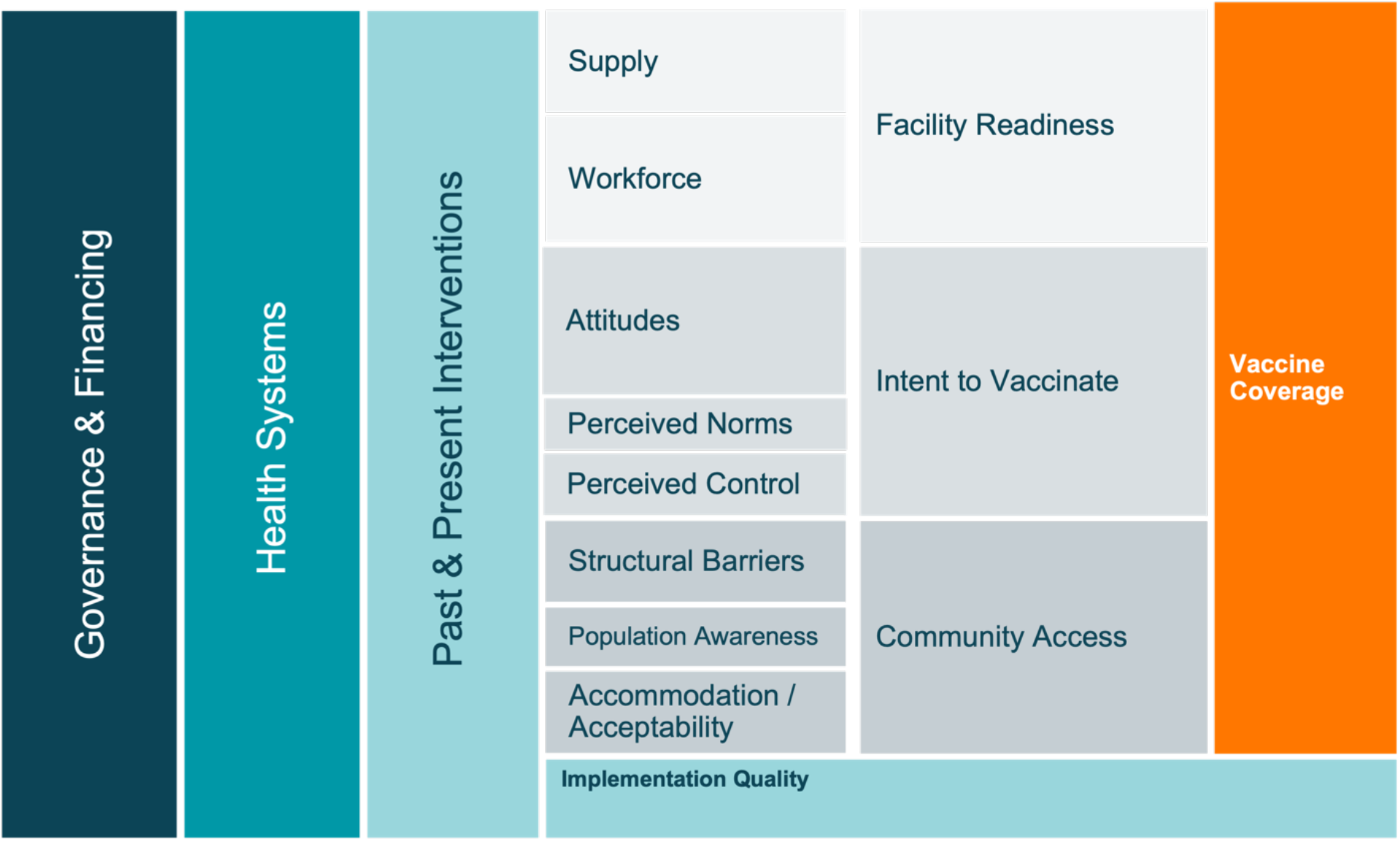
Conceptual model of the drivers of vaccine coverage.

### 2.1. Study setting

Nepal was selected as an exemplar in vaccine delivery based on DTP1 and DTP3 coverage estimates serving as proxies of the vaccine delivery system, with DTP1 serving as an indicator of access and DTP3 indicating continued use of immunization services [12].

In consultation with national stakeholders and available data, we selected three provinces as study locations while considering heterogeneity via the following characteristics: 1) geography – mountains, hills, or terai; 2) high, middling, and low performers of DTP3 coverage, as found represented across the three geographical zones of Nepal; and 3) inclusion of the capital city of Kathmandu. Province 2, Bagmati, and Gandaki Pradesh were ultimately selected with rolling 3- year averages of 76%, 90%, and 95% DTP3 coverage in 2016, respectively (Figure 2) [6, 13].

**Figure 2:**
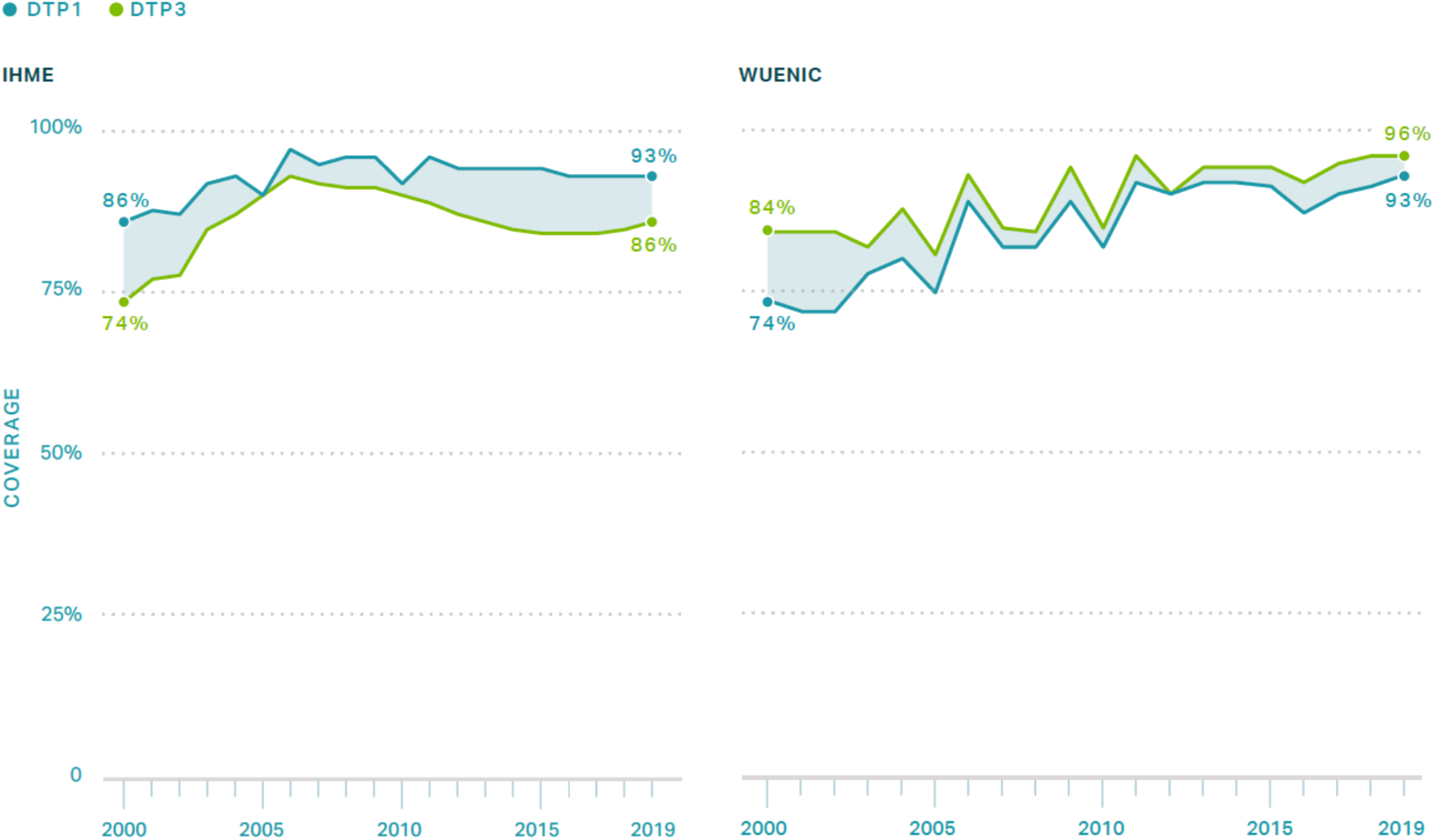
WHO UNICEF Estimates of National Immunization Coverage (WUENIC) and Institute of Health Metrics (IHME) DTP1 and DTP3 coverage of Nepal, 2000 – 2019.

**Figure 3:**
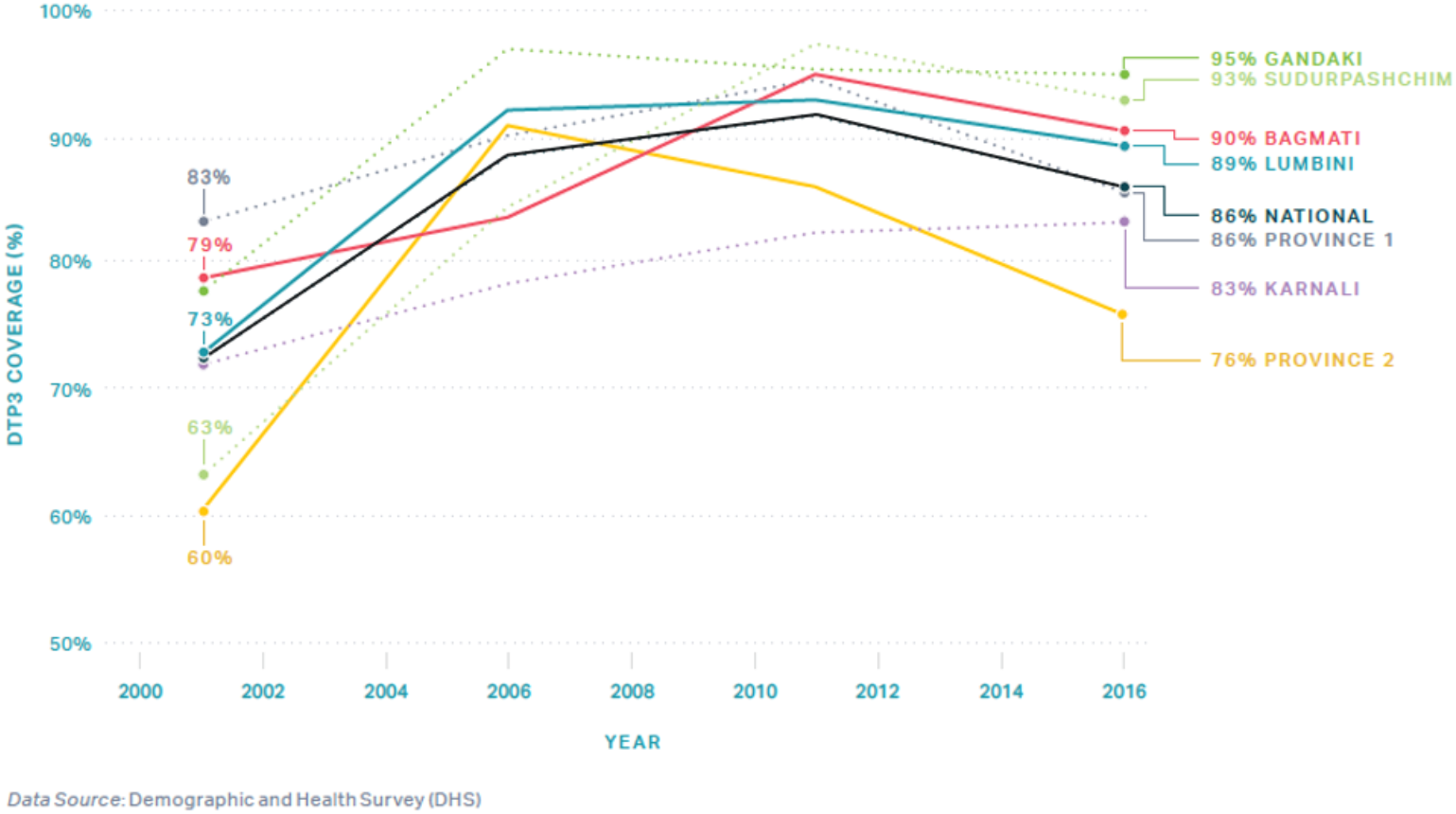
DHS DTP3 coverage of Nepal by Province, 2000 – 2016. *Note DHS data may underreport coverage estimates

We selected three districts within each province with consideration to varying DTP3 coverage and growth levels, and the re-zoning of districts that occurred in 2015 with the ratification of the new Constitution [14]. We selected the following districts: Dhanusha, Bara, and Mahottari from Province 2; Makwanpur, Dolakha, and Kathmandu from Bagmati; and Kaski, Myagdi, and Nawalparasi from Gandaki Pradesh. Health facilities were selected based on recommendations from provincial and district health directors while considering differences in DTP3 growth and coverage. One health facility was to be located near the capital of each district.

### 2.2 Quantitative data collection and analysis

Quantitative data were collected through secondary datasets, including the Ministry of Health and Population (MoHP). Data were used to estimate routine immunization coverage throughout our study period, from 2000 to 2019, and uncover trends related to improvements and sustainability. Additional analyses were conducted to identify indicators that may predict immunization coverage success among low- and lower-middle-income countries using cross-country and multi-year mixed-effects regression models to statistically test financial, development, demographic, and other country-level indicators. The results from these analyses will be presented in a forthcoming paper. Ultimately, results demonstrate that government health spending was positively associated with improvements in immunization coverage [15].

### 2.3. Qualitative data collection and analysis

Qualitative data were collected between August and November 2019 at the national, provincial, district, health facility, and community levels. The Center for Molecular Dynamics Nepal (CMDN) led data collection. The key informant interview (KII) and focus group discussion (FGD) guides – developed by the Emory research team - were informed by the Consolidated Framework for Implementation Research (CFIR) [16] and the Context and Implementation of Complex Interventions (CICI) framework [17]. Guides were translated into Nepali, with some FGD guides translated into Hindi, Maithili, and other local languages by research assistants. The interviews covered interventions, catalyzing events, and barriers and facilitators to vaccination program implementation. All interview guides were piloted before use and adjusted iteratively throughout data collection. An initial list of KIIs was developed with assistance from CMDN and MoHP officials. Snowball sampling was used to identify additional key informants. Our sampling approach aimed to include a diverse sample of participants in regard to geographic location and demographic qualities. FGDs were conducted to understand community-level factors impacting immunization coverage and to identify roles and responsibilities of frontline workers and community members within the immunization sector. Mothers, fathers, grandparents, and Female Community Health Volunteers (FCHVs) were recruited from health facility catchment areas with the assistance of local health staff. The duration of KIIs and FGDs averaged two hours. KIIs and FGDs were audio-recorded with the permission of participants. Research files, recordings, and transcriptions were de-identified and password protected. Data collection activities are summarized in Table 1.

**Table 1:** Summary of research activities, August – November 2019.

| Method | Participants | Numbers of activities | Number of participants |
| --- | --- | --- | --- |
| Key Informant Interviews | Ministry of Health and Population (MoHP), National-Level Staff | 10 | 10 |
|  | Ministry of Finance Staff | 1 | 1 |
|  | Partner Organization Staff | 8 | 8 |
|  | Provincial Health Office Staff | 5 | 5 |
|  | District Health Offices Staff | 15 | 15 |
|  | Health Post In-Charge | 10 | 10 |
|  | Immunization Officers | 3 | 3 |
|  | Cold Chain Officers | 3 | 3 |
|  | Vaccinators | 7 | 7 |
|  | Community Health Workers | 2 | 2 |
|  | Community Leaders | 15 | 15 |
| Focus Group Discussions | Female Community Health Volunteers | 9 | 60 |
|  | Mothers | 9 | 60 |
|  | Fathers | 6 | 36 |
|  | Grandmothers | 6 | 35 |
|  |  | **109** | **253** |

We applied a theory-informed thematic analysis of the transcripts to identify critical success factors. We developed a codebook through application of CFIR and CICI frameworks; the team iteratively adjusted the codebook based on emerging themes [18]. Transcripts were coded and analyzed using MaxQDA2020 software (Berlin, Germany). Relevant themes were identified, and visual tools were used to illustrate the findings. We considered setting and participant roles while identifying key points and further contextualized data using historical documents and a literature review.

### 2.4. Ethical approval

This study was considered exempt by the Institutional Review Board committee of Emory University, Atlanta, Georgia, USA (IRB00111474) and the Nepal Health Research Council (NHRC; Reg. no. 347/2019) in Kathmandu, Nepal. All participants provided written informed consent.

## 3. Results and Discussion

KIIs were conducted at national (N=19) and subnational (N=60) levels; FGDs were conducted with FCHVs (N=9), mothers (N=9), fathers (N=6),), and grandmothers (N=6) within nine districts. Table 2 provides the demographic information of 174 of 191 FGD participants.

**Table 2.** Demographic characteristics of focus group discussion participants.

| Characteristics | FCHVs<br>(n = 55) | Mothers<br>(n =54) | Fathers<br>(n=30) | Grandmothers<br>(n=35) | Total<br>(n=174) |
| --- | --- | --- | --- | --- | --- |
| Age* (range) | 47 (22-70) | 30 (19-52) | 38 (20-58) | 58 (45-75) | 42 (19-75) |
| Number of children* | 3 | 2 | 2 | 4 | 3 |
| Number of children in the home* | 3 | 2 | 2 | 3 | 3 |
| *Highest level of education\*, n (%)* |  |  |  |  |  |
| No formal schooling | 20 (37) | 18 (33) | 2 (7) | 32 (91) | 72 (42) |
| Completed primary school | 27 (50) | 26 (48) | 19 (63) | 3 (9) | 75 (43) |
| Completed secondary school | 6 (11) | 7 (13) | 5(17) | 0 (0) | 18 (10) |
| Completed post-secondary education | 1 (2) | 3 (6) | 4 (13) | 0 (0) | 8 (5) |
| *Religion\*, n (%)* |  |  |  |  |  |
| Hinduism | 46 (84) | 37 (69) | 25 (83) | 29 (83) | 137 (79) |
| Buddhism | 6 (11) | 10 (19) | 0 (0) | 0 (0) | 16 (9) |
| Islam | 2 (4) | 4 (7) | 5 (17) | 5 (14) | 16 (9) |
| Other | 1 (2) | 3 (5) | 0 (0) | 1 (3) | 5 (3) |

\*Mean taken and rounded to the nearest whole number
Note: 17 participants did not provide demographic data

We identified several government policies, health campaigns, disease outbreaks, natural disasters, and key interventions that may have affected immunization coverage in Nepal from 2000 to 2019. Figure 4 illustrates a timeline summarizing these factors as they compare to Nepal’s coverage of BCG, DTP1, DTP3, MCV1, and Pol3 vaccines [6, 19]. Funds from Gavi, the Vaccine Alliance (Gavi) are also included in this assessment.

**Figure 4.**
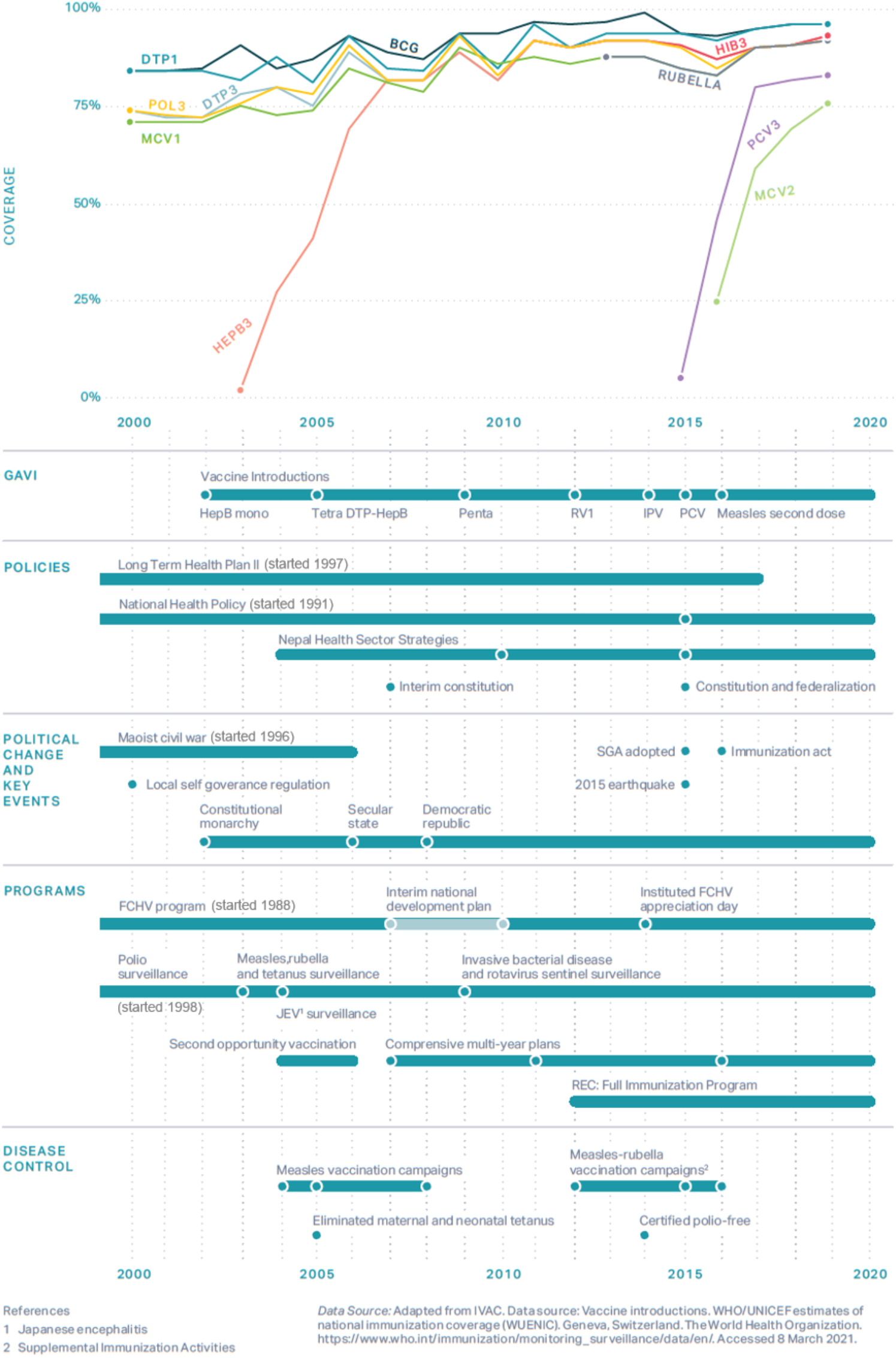
Immunization coverage with annotated events in Nepal, 2000 – 2019.

Immunization coverage had an overall increase from 2000 to 2019 with yearly fluctuations for the above-listed vaccines. Below, we describe specific strategies and policies that key informants described as success factors in Nepal.

Success factors represent both original innovations developed specifically within Nepal, in addition to adaptations of global policies to the local context. This analysis focused on how policies and statutes were formalized and operationalized to support high, sustained routine immunization coverage, as well as the means by which local context and culture shaped programming.

Our analysis led to the development of an explanatory framework that aims to categorize mechanisms of success that likely contributed to coverage improvements in Nepal. Although existing literature describes the requirements for successful vaccine delivery (Figure 1), there is a lack of evidence on how governance structures and health systems function within successful programs. Figure 5 illustrates this framework, and highlights the key drivers as they relate to Nepal’s immunization system, commitment, collaboration, awareness, and collective responsibility. We found these mechanisms contributed to successful vaccine delivery between and within the levels of implementation in Nepal. The functional definitions for these mechanisms and levels of implementation are found in Table 3.

**Table 3.**
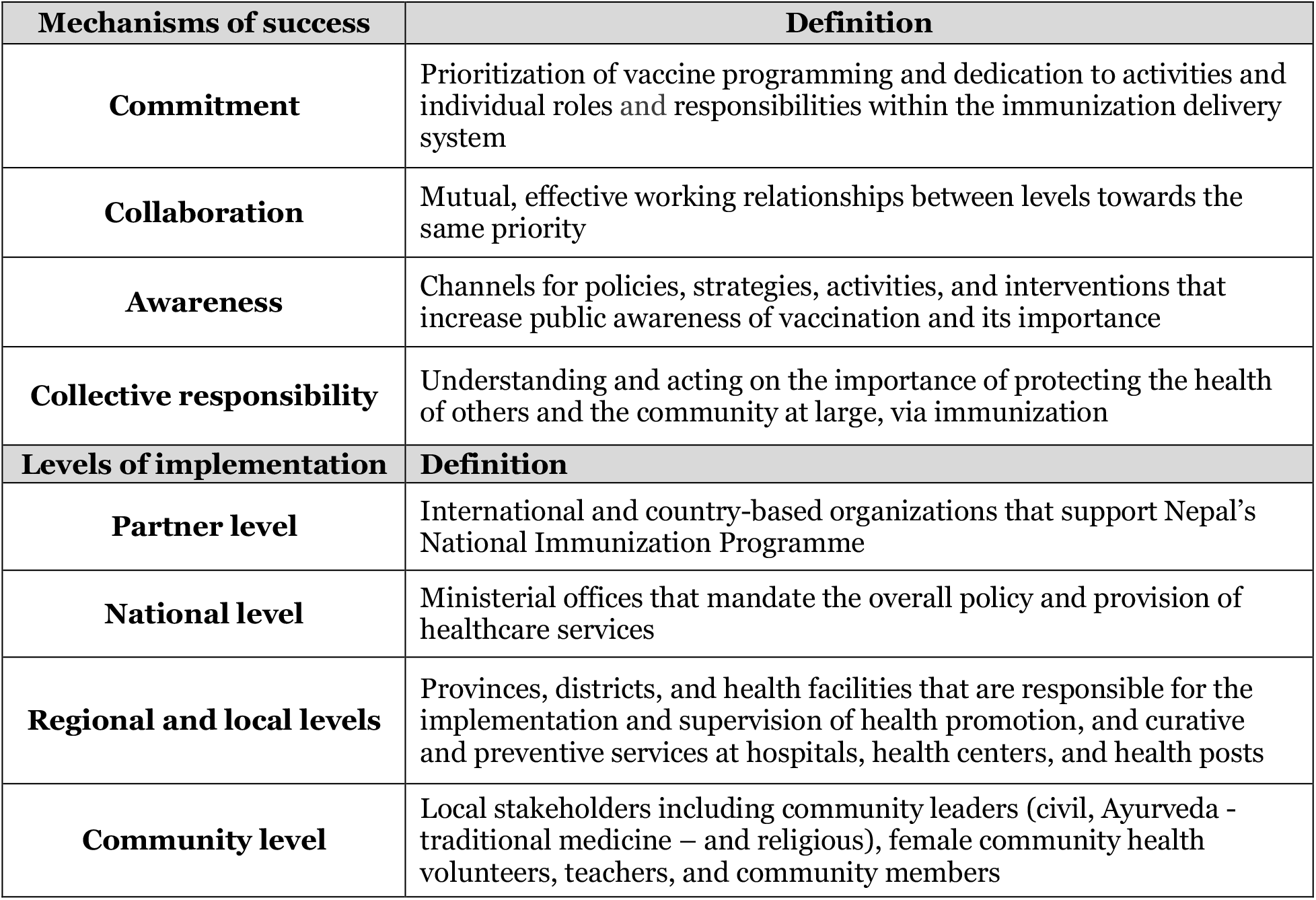
Featured functional definitions in the ‘Critical factors that contributed to high coverage of routine vaccinations in Nepal’s conceptual framework.

**Figure 5.**
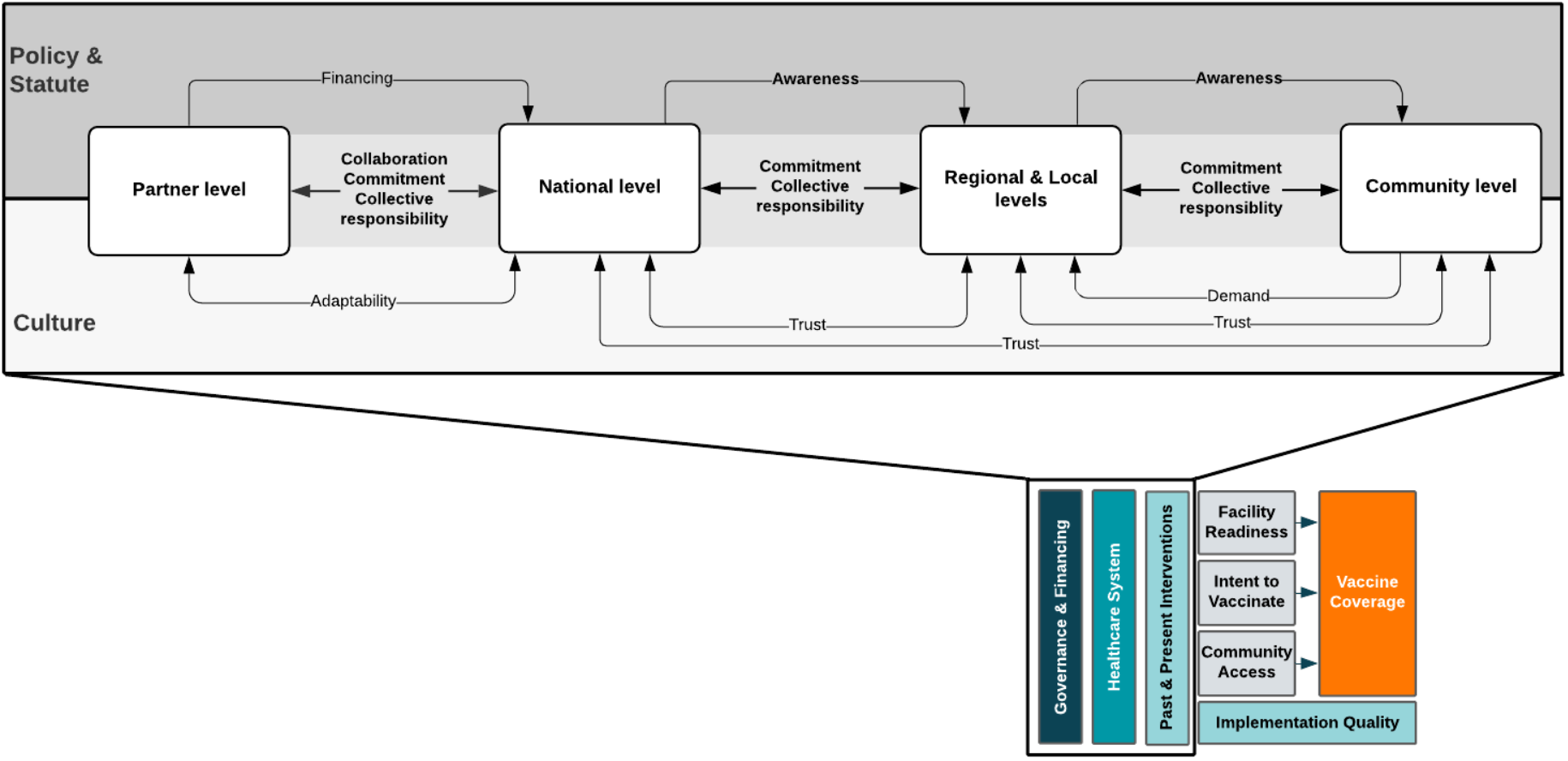
Critical factors that contributed to high coverage of routine vaccinations in Nepal. Arrows adjacent to “Policy & Statute” depict regulatory policies that Nepal operationalized into specific strategies. Arrows adjacent to “Culture” depict more informal attributes that were associated with normative and historical context.

### 3.1. Commitment mechanism

#### 3.1.1. The Constitution of Nepal codified health as a human right

Nepal’s policy environment was influenced by several declarations – the Alma-Ata declaration in 1978 and the Kathmandu Declaration in 2010. Both declarations focused on health as a human right and the expansion of primary health care services. In the late 1980s, the Nepali government recognized the need to reduce high child mortality rates. This was addressed through an increased focus on health in government decision-making, funding, and planning [20]. In 1991, the first National Health Policy was enacted with the aim of increasing primary health care services to rural and disadvantaged populations. Health posts with birthing facilities were to be established in every Village Development Committee (VDC), and FCHVs were mobilized to promote and deliver basic health services [20]. The National Health Policy of 1991 was regarded by many national-level key informants as a turning point for immunization coverage in Nepal. While not vaccine-specific, the policy affected coverage levels via overall health systems strengthening.

In 2007, Nepal’s Interim Constitution codified health as a human right, which acted as a foundation for improvements in vaccination coverage [21]. According to key informants, the recognition of vaccination as a right supported the prioritization of vaccines in policy development, motivated health workers and FCHVs, and generated community demand. The 2015 Constitution of Nepal was finalized under a newly established government, stating that: “every citizen shall have the right to free basic health services from the State, and no one shall be deprived of emergency health services.” [14] These legal documents clarified the government’s role as the duty-bearer for vaccine delivery and outreach. Although external funding and guidance contributed to the immunization program, the government remained accountable - and committed - to providing these services to all citizens. From 2000 – 2016, it is estimated that Nepal increased Government Health Spending per capita from US $4 (3 - 5) to US $9 (7 – 11); during this same period, similar LICs decreased their health spending [22].

#### 3.1.2. Legislation and budgeting ensured long-term program stability and access to vaccines

According to key informants, health policies from the national level promoted a human rights-based approach to healthcare. The emphasis on health as a fundamental right guided stakeholder efforts to improve the delivery of health services and, consequently, vaccine coverage. Key informants at the national level mentioned that the Nepal Constitution was at the center of their decision-making when adapting external policies. The government has placed an emphasis on reaching every child and is seen as the duty-bearer for health services. The government’s commitment to vaccination is further highlighted by an increased focus on lasting technical partnerships, consistent service delivery, and service delivery adaptations. Additional improvements to the health system supported the National Immunization Programme (NIP) in providing vaccines to every child, and it provided the framework for the messaging of a child’s right to be immunized.

> *“A provision was made in the constitution, and [vaccines] became a fundamental right, so the government has a responsibility to provide [vaccines]. And management of currently running programs also [focuses on] children who have missed vaccines*.*” (Former High-Ranking Official, Nepal Health Research Council)*

National policies promote equitable access to vaccines through increasing human resources, maintaining the quality of vaccine services, and focusing on sustainable health systems infrastructure. The Immunization Act, published in 2016, codified every child’s right to access quality vaccines. This act provided the foundation which continued to enhance the stability and structure of the immunization system, and consisted of comprehensive guidelines for vaccine delivery. [23] Government spending on vaccines used in the NIP was exceptionally high in 2016, with US$ 10.89 spent per birth [22, 24]. In 2015, the Nepali government spent US$ 6.29 per birth; the government spent similar (US$ 6.43 per birth) in 2017 [15]. Informants at all levels of the system commented on Nepal’s continued commitment to health and vaccination over the years. Steady increases in spending over time demonstrate Nepal’s commitment to the NIP – for example, the government spending on RI vaccines per birth in 2006 was only US$ 1.18 [15].

The Immunization Act also created a dedicated national Immunization Fund to allocate money for the NIP when Nepal is no longer receiving external support [25-27]. The Immunization Fund was created with support from the Sabin Institute – a global vaccination development non-profit –and is a public-private partnership between the Nepali government and Sabin [25, 28]. It acts as a savings account to prepare for the expected transition off Gavi support and ensures Nepal has the ability to fund immunization programming.

### 3.2. Collaboration mechanism

#### 3.2.1. The partnership between the MoHP and WHO supported national-level system improvements

Effective collaboration was fostered through key partnerships between external partners and the MoHP. These partners include, but are not limited to, WHO, UNICEF, Gavi, Rotary International, and the Global Polio Eradication Initiative (GPEI). Key informants from all levels reported high levels of trust, mutual respect, strong coordination and collaboration, and an agreed-upon division of labor among the MoHP and external partners. Key responsibilities within the immunization sector are divided as such: UNICEF works on communication materials, trainings, and supply chain maintenance; WHO position papers are used for vaccine recommendations; and the MoHP oversees all policy development, implementation, and coordination.

The partnership between the WHO Immunization Preventable Diseases (IPD) program and the MoHP started with polio surveillance over 20 years ago. In 1998, WHO started the Polio Eradication Nepal (PEN) office, and supported duties associated with the surveillance system.^[29]^ Due to dwindling cases of polio, the PEN office was at risk of closure. The MoHP did not want to lose this partnership, and in 2005, PEN was renamed to the IPD office; their scope expanded to support the MoHP on all vaccine-preventable diseases (VPDs) in addition to epidemic and outbreak surveillance activities [29]. Participants from both the MoHP and WHO noted the importance of WHO-IPD, with a former MoHP official noting that IPD is the “backbone of the [surveillance] system,” and another stating they provide the ministry with “maximum support.”

WHO-PEN, and then WHO-IPD, built the capacity of the surveillance system gradually. Each new indicator was added to the already existing system building on top of preexisting infrastructure. Collaboration from partners, especially WHO-IPD, was crucial to this growth. The resilience and flexibility needed to maintain and grow the system speak to the investment in the partnership. The WHO-IPD office is seen as integral to the immunization system by participants both inside and outside the ministry. The MoHP and WHO-IPD work on routine immunization programming as partners, and share resources and data as needed to reach common goals.

The Global Polio Eradication Initiative (GPEI) has provided the bulk of the funding for this programming; GPEI continues to fund the WHO-IPD office even though its activities are no longer heavily focused on polio elimination. Funds are not siloed, and no distinction is made between polio activities and routine immunization activities. The notion that individual diseases do not exist in a vacuum is critical to the success of the immunization programming and to their collaboration.

> *“We have gradually taken upon ourselves, thanks to some donor support, [a role] beyond GPEI. GPEI still remains the mainstay of our support. But beyond GPEI, GAVI, CDC, USAID, others, UN Foundation, etc., we have morphed into a team that is supporting increasingly not only measles immunization and rubella, but also routine immunization.” (External Development Partner)*

### 3.3. Awareness mechanism

#### 3.3.1. Pro-vaccine messaging supports public awareness through multiple media outlets that are tailored to local needs

The MoHP allocates media engagement responsibility to a governmental agency, the National Health Education Information and Communication Center (NHEICC). The NHEICC coordinates with MoHP departments, external partners, and media agencies - including newspapers, radio, and television networks - to craft health-related messaging, and utilizes social media platforms to relay information. Media is used to spread general information about immunizations, in addition to promotion of routine vaccination days, National Immunization Days, and the introduction of new vaccines. The government is responsible for providing accurate and timely information to the media, and verifies and corrects messaging as needed.

> *“We are closely related to the media since the beginning of time. We [summarize] our activities and progress to the media – not only at the national level but also in lower levels. After we worked with the media, they gave priority to the immunization program.” (Former Official, World Health Organization)*

Most key informants, including health workers and parents, stated that media is a trusted source of information. However, that might not be true across all communities in Nepal. Previous research states that communities in the Terai region are less likely to trust messaging from the media, and more likely to trust community members - including FCHVs [30].

Health workers adapted messages from the national level to meet context-specific needs. Key informants from all three provinces mentioned budgets allocated for media exposure. However, media cooperatives also seemed to offer free services when funding was unavailable.

> *“The media must want to support the immunization program in order to help the children, without thinking to disseminate the news only for money. They can do this by coordinating with the health office. We provide money to the media in order to disseminate the news*…*” (Cold Chain Officer, Mahottari, Province 2)*

Key informants referenced frequent community-level review meetings to discuss how to address challenges, such as misinformation or poor uptake. Leadership within the municipality and Village Development Committee (VDC) offices - the lowest administrative levels in Nepal - may offer guidance and technical support for community workers adapting messaging or outreach activities. Ultimately, the impetus for adaptations came from within the communities where workers and volunteers could relate to any cultural, social, and geographical challenges faced by friends, family, and neighbors.

#### 3.3.2. Consistent community engagement and education generated strong demand for vaccines

Public awareness for vaccines was sustained through extensive community engagement led by the national level MoHP and implemented through community health workers and volunteers, which was crucial to support the community-centered social movement for health in Nepal. Community-led leadership and committees - including traditional leaders, government officials, volunteers, mothers, and teachers - were formed by the MoHP to address high childhood mortality. Community-based groups meet with district-level health officers to discuss the state of vaccination, challenges, and solutions within their municipality. These community-led groups are involved with immunization programming, including promotion, service delivery, and targeted outreach. Committees are comprised of members from a variety of religious, ethnic, and socioeconomic groups to connect with hard-to-reach communities and ensure rumors about vaccines are dismissed. Involving diverse stakeholders allowed for tailored messaging and outreach by leaders at the ward or village level depending on contextual factors including culture, religion, and geography.

> *“The vaccination programs are not differentiated based on any tradition, culture, or language. Whether the people follow any religion or are at any caste, the vaccination awareness and services are [provided] to everyone.” (Immunization Supervisor, Dhanusha, Province 2)*

Community leaders in all provinces demonstrated a significant commitment to their communities’ health through conducting extensive household visits, reminding parents to bring their children to vaccination days, and sometimes offering transport to vaccination sites.

> *“We gave them the example of well-fertilized and unfertilized crops, and then showed them the pictures of vaccinated and unvaccinated children. Then finally we were able to convince them that vaccination is an essential thing for all the children.” (Community Leader, Nawalparasi, Gandaki Pradesh)*

The importance of vaccination is taught to school children as part of their standard curriculum. The education system plan of 1971 made health a compulsory subject for all grade levels and was created to influence the health knowledge, attitudes, and practices of students. The Ministry of Education developed a standard curriculum for “Health Education”, and only educators approved by the MoHP can teach this curriculum, which covers a variety of health topics, including communicable diseases; vaccinations; personal hygiene, water and sanitation; nutrition; safety; and first aid.[31, 32] School teachers were mobilized to strengthen immunization coverage and were involved in immunization activities.[33] Additionally, teachers inform their students about the date and place of vaccination sessions in their respective VDCs, and were involved in checking the immunization status of at-risk populations.[34]

> *“If the information [is communicated] by teachers, it will surely be obeyed by the local people, and if we go instead of teachers, it will require more time.” (University Personnel, Tribhuwan University Institute of Medicine)*

Improvements in general education and literacy rates among mothers increased alongside childhood immunization coverage. There was an overall increase in literacy of mothers with 1 year old children from 35% (32% - 37%) in 2001 to 67% (63% - 70%) in 2016. During that time period, there was a higher rate of literacy seen in mothers of 1 year old children who did not miss any vaccines. In 2001, those mothers had a literacy rate of 45% (41 - 48%) compared to those mothers whose children had missed at least one vaccine with a literacy rate of 15% (12-19%). Both groups increased by 2016; however, the mothers with fully vaccinated children maintained a higher literacy rate (71% [67-74%], 53% [46 – 61%] respectively) [35].

#### 3.3.3 FCHVs were trusted advisors and critical messengers who promote health and access to vaccines in their communities

FCHVs implement public awareness programs at the community level, and are critical to healthcare in Nepal. Established in 1988, FCHVs provide essential services: promotion of and education on preventative health care, including vaccines and vaccination services, and raising public awareness [36]. Providing support to the NIP was added as a FCHV responsibility in the National Health Policy of 1991, including the use of defaulter training to identify mothers not accessing vaccination services, visiting individual households to address misinformation, and addressing caregivers’ concerns. Most importantly, FCHVs taught caregivers about vaccine-preventable diseases, and in turn, caregivers began to prioritize routine vaccines for their children.

FCHVs are married or widowed women who are required to work in the communities they reside in; mothers reported that this generates trust, and promotes culturally appropriate messaging. To address different stakeholders, messaging is adapted to be context specific. In Mother’s Groups, health and immunization education was presented in monthly meetings; content was selected and tailored depending on literacy, misinformation and cultural norms. In women’s savings groups, FCVHs emphasized the link between children’s health and future educational and occupational opportunities. FCHVs also approached caregivers individually, discussing potential barriers to immunization, addressing specific information caregivers lacked, and encouraging immunization access. As FCHVs were embedded in the communities where they conducted outreach, their messaging incorporated differences in religion; tribe or caste; ethnicity; and language. Community recruitment increased outreach in remote areas.

Mothers’ Health groups (MHGs or “Ama Samuha”), one of the oldest civil society groups in Nepal, collaborate with FCHVs to deliver community education. MHGs are formed with the involvement of the community, local health institutions, and local governments, and consist of “all interested mothers of reproductive age” [36]. Key informants in all districts praised their nationwide active involvement in community health. FCHVs organize these monthly informational meetings with MHGs to discuss maternal and child health (MCH) topics, including immunizations. FCHVs, CHWs, leaders for local women’s groups, teachers, and regional experts may facilitate. MHGs are essential to community peer-to-peer education; group members are expected to share information with other caregivers. The MoHP recently emphasized recruiting MHG members from marginalized groups to improve equity, awareness building and community trust [36]. Mothers’ group meetings occur in every province in Nepal and in most communities; the 2019 DOHS Annual Report stated that there is a consistent increase in MHGs in every district [37].

> *“Basically, mothers’ groups are responsible to give education and information to the parents*… *If a mother knows about the immunization [days], then the mother takes her baby in any condition or situation. FCHVs and Mothers’ groups provide information regarding immunization [days] or the regular immunization program. New vaccines need to [be] discussed with FCHVs and mothers’ groups because unless they support and are involved in the program, [the introduction of the new vaccine] will not be successful because [FCHVs and mothers] are the ones who build awareness and give knowledge and information to the people at the grassroots level.” (Director General, Department of Health Services in the Ministry of Health and Population)*

### 3.4. Collective responsibility mechanism

#### 3.4.1. Cultural norms of collective responsibility foster community engagement and ownership of vaccine programming

Empathy is a widely accepted cultural value according to our key informants, and community needs are regarded as more important than individual needs. These norms contributed to the collective responsibility and ownership of local vaccination coverage.

> *“Health is everybody’s responsibility - it is incomplete even without support of just one small community. What I think is that until there is a healthy person, a healthy community, and a healthy family, there is no healthy nation.” (District Health Officer, Bara, Province 2)*

Government workers, external donor organizations, non-profit organizations, and parents were all dedicated to the common goal of increasing vaccine coverage through distinct responsibilities. As it is the right of every child to receive vaccines, it is the responsibility of health staff to provide vaccines, the responsibility of the state to provide health staff with the resources required, and the responsibility of parents to take their children to be vaccinated. This collaboration is fostered by the government through transparency in decision making and vaccine effectiveness; consistent and reliable service delivery; communication of government policies to the communities; and shared cultural values of altruism and collective responsibility. Key informants identified these partnerships as essential components of vaccination programming.

> *“In immunization, there are three main stakeholders: One is the government; second is UNICEF and WHO - they are donors - and last is the community. These three are responsible for improvements in the immunization coverage.” (Personnel, UNICEF)*

Local government officials noted that recognition of health as a right of all citizens motivated participation in the vaccination program. FGDs with FCHVs revealed that a social obligation to their communities, especially the children, motivated their efforts to increase vaccination coverage. FCHVs volunteer their time to ensure community members are knowledgeable of their right to receive vaccines, and they engage directly with the communities through door-to-door outreach to parents. Their information dissemination and continuous education work to increase public awareness. A structured, consistent, and reliable vaccine delivery system, with vaccination days on the same date and at the same site every month, aligns with this continuous education. In rare circumstances of vaccine shortages, parents will ask vaccinators, FCHVs, and other MoHP clinic staff why their children cannot receive vaccinations and when they can expect these services again; communities were disappointed that their expectations were not met.

> *“People in the community think that immunization is very important. They think that it is their right. The parents call to ask when they should bring their child for vaccination.” (Health Post in Charge, Kaski, Gandaki Pradesh)*

### 3.5. Implications for strategic planning in immunization programming

We conducted a theory-and evidence-based mixed methods investigation into the historic drivers of how and why Nepal was able to achieve catalytic growth in early childhood vaccine coverage. Rather than identifying what interventions targeted intent to vaccinate, community demand, and facility readiness, our data support the need to better understand core governance structures and functions with regards to commitment, community ownership, and data transparency and use. Commitment, collaboration, awareness, and collective responsibility at multiple levels of the health system was crucial for the success in Nepal, a finding that applies likely beyond vaccine delivery to the entire health system and suggests the importance of more systemic assessment of health delivery.

Nepal’s success in increasing routine immunization coverage heavily relied on its highly collaborative network of partners. Nepali government agencies leveraged partnerships in order to strengthen the overall vaccine system, utilize unique and diverse skills from other stakeholders, and foster prioritization of vaccination. International partner organizations working in Nepal are thoroughly invested in the success of the NIP, and work with the government as part of a team. Collaboration between the MoHP and WHO-IPD led to significant improvements in the country’s surveillance system. The government also worked with the private sector; coordination between the NHEICC and media agencies (e.g., radio stations) supported increases in public awareness. Across the system, leveraging partnerships proved important to successful vaccine delivery.

Structured policies, often taken from global recommendations, proved important to initially standardize and guide implementation. These policies are beneficial for much of vaccine delivery, but true success comes from adaptation at the community level. Community input and guidance allow for policy to be informed by the context. Engaging the community in these efforts created the best environment for uptake, and several key informants believed the high vaccine coverage stemmed from involved and aware citizens. Community involvement was crucial not only for local adaptation but for demand generation. FCHVs and health workers helped to foster an environment where everyone was involved in health and health decision-making, including men, women, children, and elders. This demand environment and the general public awareness were supported through cultural norms and were eventually codified – showing citizens that it was their legal right to be vaccinated.

The legal framework set in place by Nepal created a policy environment focused on immunizations. With the National Health Policy of 1991, health became a major focus of the government, allowing for the initial structure of the NIP. Prioritizing health, and the use of cultural norms led to the codification of health as a human right in the 2007 interim constitution. This prioritization was further specified in the Immunization Act, which laid out regulations for vaccination programming. Codifying immunization cemented it as a high priority of the government both legally and culturally, creating room for the program to expand and improve.

Findings from this research highlight the need for programmers and policy makers to better understand the strengths and limitations of the underlying governance structures in terms of fostering commitment, collaboration, awareness, and collective responsibility at national, sub-national, and local levels. There is no panacea to apply these lessons; however, our approach to understanding underlying contextual and program delivery processes may have implications on vaccine policy, programming, and investments in other low- and middle-income countries. Though some of the strategies described in this paper may be unique to Nepal, many of the highlighted success factors may have salience in other settings or apply to other health systems for a horizontal approach to healthcare.

## 4. Limitations

This study has several limitations. First, we focused on Nepal as a positive deviant in vaccine delivery but were unable to carry out a similar analysis in a non-exemplary country to compare immunization coverage. Second, the research tools focused on eliciting critical success factors and did not probe on interventions or policies that were unsuccessful. Third, using qualitative methods to understand historical events was challenging; interviewees often spoke about current experiences rather than discussing historical factors. However, research assistants probed respondents to reflect on longitudinal changes in the immunization program. Fourth, while qualitative findings were based on a breadth of stakeholders at different levels, we were limited by the samples we received. Lastly, some policy documents and datasets had limited accessibility by time, language, and paper over electronic records, even with the assistance of our in-country partners.

## 5. Conclusion

Data from Nepal suggest that the success of vaccine systems was supported by consistent and reliable commitment, collaboration, awareness, and collective responsibility between the government, the community, and partner networks. The involvement of community members in vaccination programming was essential to the uptake of vaccine services and contributed to high coverage. These networks were strengthened through a collective dedication to vaccination programming and a universal belief in health as a human right.

## Data Availability

De-identified data are available upon request.

## Acknowledgements

Center for Molecular Dynamics Nepal (CMDN) was a key partner in this study. Technical support and guidance were provided by Sarah Chesemore, Anna Rapp, Tove Ryman, and Ethan Wong from the Bill & Melinda Gates Foundation and Kate Buellesbach, Nathaniel Gerthe, Gloria Ikilezi, Angela Hwang, Caitlyn Mason, David Phillips, Oliver Rothschild, and Jordan-Tate Thomas from Gates Ventures. The Vaccine Exemplars Research Advisory Group included Agnes Binagwaho, Laura Craw, Carolina Danovaro, Anuradha Gupta, Heidi Larson, Penelope Masumbu, Kate O’Brien, Helen Rees, Lora Shimp, and Aaron Wallace. We gratefully acknowledge the participants from Province 2, Bagmati, and Gandaki Pradesh who gave their time and insights to help us better understand Nepal’s vaccine delivery system, along with the Ministry of Health and Population, Department of Health Services, and the Family Welfare Division of Nepal.

## Data statement

De-identified data are available upon request.

